# Characterizing an epidemiological geography of the United States: influenza as a case study

**DOI:** 10.1101/2021.02.24.21252361

**Authors:** Grant E. Rosensteel, Elizabeth C. Lee, Vittoria Colizza, Shweta Bansal

## Abstract

The prediction, prevention, and management of infectious diseases in the United States is either geographically homogeneous or is coordinated through ad-hoc administrative regions, ignoring the intense spatio-temporal heterogeneity displayed by most outbreaks. Using influenza as a case study, we characterize a regionalization of the United States. Based on influenza time series constructed from fine-scale insurance claims data from 2002-2009, we apply a complex network approach to characterize regions of the U.S. which experience comparable influenza dynamics. Our results identify three to five epidemiologically distinct regions for each flu season, with all locations within each region experiencing synchronous epidemics, and with an average of a two week delay in peak timing between regions. We find that there is significant heterogeneity across seasons in the identity of the regions and the relative timing across regions, making predictability from one season to the next challenging. Within a given season, however, our approach shows the potential to inform on the shaping of regions over time, to improve resources mobilization and targeted communication. Our epidemiologically-driven regionalization approach could allow for disease monitoring and control based on epidemiological risk rather than geopolitical boundaries, and provides a tractable public health approach to account for vast heterogeneity that exists in respiratory disease dynamics.

## 1 Introduction

The spatio-temporal dynamics of infectious diseases are complex to interpret, but are key to the success of public health responses. Seasonal influenza, for example, exhibits variability in onset, peak time, duration, and geographic distribution between seasons, and is thought to be affected by a number of environmental, socio-demographic, and biological factors [1, 2, 3, 4, 5]. In the United States, surveillance, vaccination policies, and resource allocation for influenza are managed at the level of ad-hoc health administration mega-regions (each consisting of multiple U.S. states) [6]. Current sentinel surveillance provides a region-level situational awareness on seasonal influenza epidemics, however it provides limited information due to its aggregate nature [1]. Basing surveillance, prevention, and control efforts on large geographies grounded on administrative boundaries fails to capture the underlying spatial variation in epidemiological dynamics of infectious disease.

An alternative to existing strategies is *regionalization* which groups localities together based on epidemiological risk rather than traditional geopolitical boundaries. Operationally, regionalization has been leveraged in public health to improve coordination, standardization, and centralization of public health services to maximize constrained resources and minimize poor outcomes [7]. However, identifying regions in a regionalization strategy can be complex, particularly for infectious diseases, given the different and sometimes opposing goals: epidemiological heterogeneities may exist at very small scales, while large spatial regions may be required for efficient public health management. Successful examples of infectious disease regionalization can be found in animal health management efforts with the Designated Surveillance Area plan for brucellosis in Yellowstone National Park [8] and the bovine tuberculosis zones in Michigan and Minnesota [9]. However, such efforts remain on a small scale, and implementing nationwide regionalization requires an empirically-driven analytical tool which selects regions based on epidemiological dynamics.

The analysis of complex networks may provide an analytical approach to regionalization. If network nodes represent individual geographical units (e.g. counties), and edges represent a functional similarity of epidemic time series between pairs of counties; the analysis of the large-scale network structure properties would allow for a holistic study of the connectivity and make it possible to optimize regions from a global perspective. Community structure detection, also known as graph partitioning, [10, 11, 12], is a method of large-scale network structure analysis that aims to identify *communities* based simply on the information encoded in the network structure, where a community is defined as a set of nodes more densely connected to each other than the rest of the network. In general, communities are of intrinsic interest because they correspond to functional units within the network that can be identified without any extrinsic knowledge about the node characteristics. Epidemiologically, such network communities can correspond to spatial regions composed of locations that have similar disease dynamics, thus creating an “epidemiological geography.”.

An analytical regionalization approach necessitates a high-resolution source of epidemiological data that can distinguish transmission dynamics at smaller spatial scales. One such source of disease data with high geographic resoluation is medical claims; in the United States, medical claims for influenza-like illness (ILI) have been validated at multiple spatial scales to sentinel surveillance data reported by the roughly 3000 physicians participating in the CDC system ILINet [13], and have identified population-level associations with factors thought to drive influenza transmission [1].

Previous work also hints at ecological mechanisms that may contribute to natural influenza regions. In the United States, while influenza season introductions may be linked to air travel, seasonal influenza spreads spatially in hierarchical wave-like patterns [14] through local travel patterns [2, 5, 15, 4]. Consideration of age patterns is also thought to be an important component to understanding the spatial patterns of transmission. Within influenza seasons and pandemics, school-aged children are thought to drive local influenza transmission because of their large number of contacts, while adults are thought to seed influenza in different locations owing to their global mobility [16, 17, 18, 19].

In this work, we leverage a large-scale fine-grain data source on influenza incidence to characterize an epidemiological geography for the United States composed of regions with similar epidemiological dynamics. We hypothesize that such a regionalization will illustrate the spatio-temporal dynamics of influenza epidemiology, and elucidate variation in dynamics across seasons. We also hypothesize that epidemiologically-driven influenza regions will be spatially contiguous overall, and that the regions defined by adult influenza dynamics will be more spatially dispersed than the regions describing child dynamics. This quantitative regionalization approach can be used to inform spatial public health policy at the federal and state levels for identifying optimal sentinel surveillance locations, resource allocation, and targeting of interventions such as vaccination.

## 2 Methods

We adapt network-based time series clustering methods to identify regions with similar ILI dynamics across the United States (Figure 1) [20, 21]. We define a spatial network where nodes represent U.S. counties, and edges represent similarity in disease dynamics between each pair of locations. We analyze the spatial structure of this network, and then perform community structure detection to identify groups of locations with homogeneous epidemiological dynamics, taking into account practical limitations. Finally, we consider predictive models on the results to evaluate the potential for nowcasting or forecasting epidemiological dynamics based on region membership or regional membership based on epidemiological dynamics or covariates.

**Figure 1:**
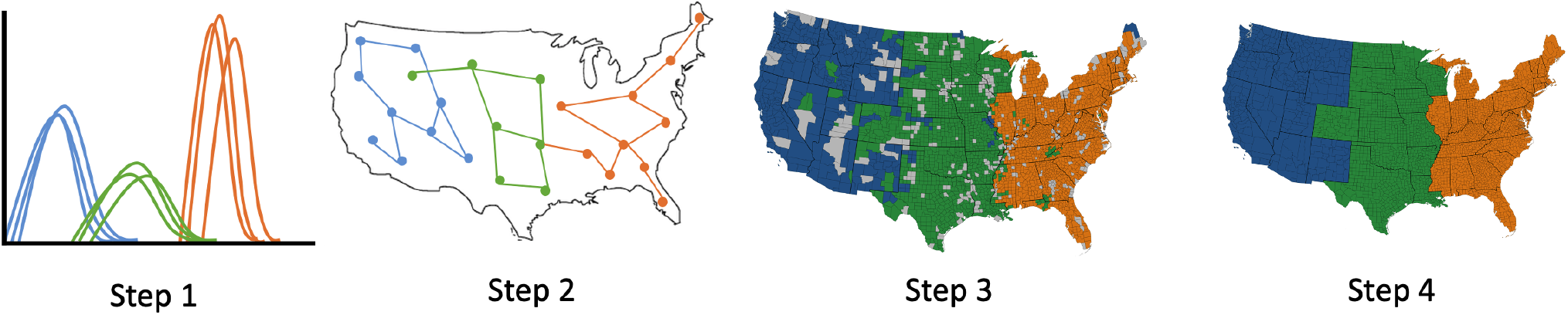
Methods Overview. Step 1: Isolate incidence time series for the disease in question, at a fine spatial resolution. Step 2: Generate a similarity network in which edges link locations that experience similar epidemics. Step 3: Perform community structure detection on the similarity network to identify geographic regions with homogeneous epidemiological dynamics. Step 4: Increase spatial contiguity of geographic regions for ease of public health implementation.

### 2.1 Influenza Incidence Data

Weekly visits for influenza-like illness (ILI) and visits for any diagnosis were obtained from a records-level database of US medical claims managed by SDI Health (now IQVIA) from October 2002 to April 2009 and aggregated to three-digit patient U.S. zipcode prefixes (zip3s). ILI was defined with International Classification of Diseases, Ninth Revision (ICD-9) codes for: direct mention of influenza, fever combined with respiratory symptoms or febrile viral illness, or prescription of oseltamivir. Previous work has compared these data to U.S. CDC sentinel surveillance and other sources [13] and discusses the data coverage, data processing, and its characterization of U.S. ILI spatiotemporal dynamics in greater detail [1, 2]. Our data represented roughly 24% of visits for any diagnosis from approximately 37% of all health care providers across 95% of U.S. counties during influenza season months, averaged over the years in our study period. We use ILI data for the total population (all age groups) in each U.S. county, as well as data specific to ILI among school-age children (aged 5-19 years), and adults (aged 20-69 years).

As described in detail elsewhere [1], we defined a county-level time series of weekly *ILI ratio* as the percentage of visits for any diagnosis that were due to ILI, as it accounts for seasonal variation in total underlying visits to healthcare providers. We detrended each county-level time series of ILI ratio and fit a linear regression model with annual harmonic terms and a time-trend to non-flu period weeks. Counties had “epidemics” in a given flu season if at least two consecutive weeks of detrended ILI observations exceeded the epidemic threshold (defined as the upper bound of the 95% confidence interval for the linear model prediction). These detrended epidemic time series were then z-normalized to put them on the same scale.

### 2.2 Constructing Time Series Similarity Networks

To generate time series similarity networks, we calculated the pairwise similarity between z-normalized seasonal epidemic time series for each pair of counties, *i* and *j* using the Pearson correlation coefficient (*ρ*_*ij*_). (We note that this is equivalent to the Euclidean distance for normalized time series, [22]). Each U.S. county was specified as a node in the epidemic similarity network, and pairs of counties, *i* and *j*, were connected via an edge if *ρ*_*ij*_ ≥ 0.8. Edges were weighted with *ρ*_*ij*_. We selected a similarity threshold (0.8) after performing a sensitivity analysis to optimize epidemiological utility. (For more information on our inference of networks from epidemic time series data, please see the supplement.) We construct such networks from ILI data for the entire population, and separately for the adult and child populations.

### 2.3 Analyzing Time Series Similarity Networks

We analyze the spatially-embedded network of epidemic similarity by measuring the spatial and network structure of the inferred networks. In particular, we measure the degree distribution of the network. The degree represents the number of counties that share similar epidemic dynamics with a given node, thus the degree distribution, *p*_*k*_ summarizes the distribution of connectivity at the county level. We fit the observed degree distribution to a power law distribution (*p*_*k*_ = *ck*^*-θ*^).

We measure spatial dispersion, the average distance (in kilometers) of the neighbors (county nodes with which a given county node shares edges) of each county, as a summary statistic of the spatial structure of the network. We hypothesize that the spatial dispersion of the epidemic similarity network for the total population will differ from the ones for other demographic groups, particularly that children will have a lower spatial dispersion on average than adults or the total population.

### 2.4 Clustering Time Series: Community Detection

We propose the use of complex network community structure detection to provide a quantitative approach to public health regionalization efforts. This method will effectively cluster nodes (counties) into groups of nodes (geographical regions) that are more connected within than between. The resulting clustering will thus represent a regionalization of the U.S. in which regions consist of counties that have more similar influenza epidemics to each other than to other regions. One benefit of the network-based community detection approach over traditional clustering methods is that community detection does not require user specification of the number of clusters (regions, in this case); instead the number of clusters emerge organically from the data connectivity [23].

For community detection, we use the Louvain method [24], a multiscale method in which modularity is first optimized using a greedy local algorithm. An initial application of the algorithm to our county-level network produces a “supernetwork” — a network where nodes represent the algorithm-identified communities — that maximizes modularity, a network-level metric of community detection [25]. The algorithm is applied iteratively on each successive supernetwork until no further improvements in modularity are possible. This method has become widely used due to its computational efficiency and high quality results [26, 27, 28]. We perform the Louvain algorithm on the similarity network with edge weights (i.e. time series correlations) using a igraph implementation in *Python* [29]. We also validated our results using two networkx implementations [30, 31]. (More information in supplement).

#### 2.4.1 Partition quality

We measured the quality of a community structure partition using two measures: (a) *relative modularity*, which can be calculated as the fraction of within-community edges beyond a random expectation [25]; to allow for the comparison of modularity values between networks of different sizes, we normalize the modularity by the maximum modularity possible for that network [32]; (b) *silhouette*, which is a measure of how similar a network node is to its own community compared to other communities [33]. *Normalized mutual information* (NMI) was used to compare pairs of community structure partitions [34]. To measure the cohesion of each region, we also measure the *intraregion connectivity*, as the proportion of edges in a region that are within the region, and the *intraregion similarity*, as the average time series similarity within a region.

#### 2.4.2 Robustness

We performed a robustness assessment of the community structure using a set of “bootstrap networks”, {*B*_*i*_} where *i* = 1, .., 10. For each bootstrap network, network edges were preserved from the original network, but for each edge, a new edge weight was drawn from a Poisson distribution with mean *w*_*ij*_, the original edge weight (i.e. time series correlation). The community structure algorithm was performed on each bootstrap network. A consensus value, *β*_*i*_, was then calculated as the sum of the NMIs between the community structure partition of bootstrap network *B*_*i*_ and all other bootstrap networks. The partition with the largest consensus value was defined as the robust community structure partition.

#### 2.4.3 Feasibility

The partitioning identified by our community structure algorithm, while theoretically optimal, may be challenging for public health implementation as it may contain a number of small regions, and each region may not be spatially contiguous. We thus introduce methods to make the epidemiological regionalization feasible for public health implementation. First, we eliminated small regions, *≤* 94 counties in size (3% of all US counties). Second, we developed an algorithm to increase feasibility by eliminating counties with missing communities and island communities that are disconnected (geographically) from other parts of the community. To achieve this, we first define the nearest neighbor network, *G*_*nn*_, in which an edge exists between a pair of nodes if they are spatial neighbors, that is, they share a boundary in geographical space. For each community, we consider the nearest neighbor subgraph of *G*_*nn*_ made up by the nodes in that community; we then identify small (i.e. fewer than four nodes) disconnected components and merge them with the most common neighboring community.

To measure the success of our algorithm, we define *spatial contiguity* is defined as the mean reachability (the fraction of node pairs that have a path from one to another) of all community subgraphs of the nearest neighbor graph, *G*_*nn*_. Thus, spatial contiguity requires that all parts of a district be physically connected; practically speaking, it is a region in which one can travel to all parts of without entering a different region.

#### 2.4.4 Validation

We validated the community structure partition results methodologically and epidemiologically. To validate our results methodologically, we performed a traditional hierarchical clustering method on the normalized epidemic time series (implemented using the scipy.cluster module in *Python*) on our data. We compared our community structure partition results to the hierarchical clustering results using NMI.

To validate our results epidemiologically, we focused on two metrics of public health importance: the influenza season onset time, defined as the first week above the epidemic threshold; and the peak time, the week during which the largest incidence was experienced during the outbreak. We used Welch’s unequal variance t-test to measure if the onset times and peak times of different communities or regions identified within our community structure partition were statistically different.

### 2.5 Predictive Methods

#### 2.5.1 Prediction of Time Series

To measure the potential of epidemic dynamics in one region to forecast the epidemic dynamics in another region during the same season, we measure the Granger causality between the epidemic time series from different regions. We first verify that the time series have order of integration of one, meaning that the differenced time series with a lag of one is stationary. Then, we measure the Granger causality using the F distribution between the average epidemic time series for region *i* and *j*, where the epidemic in region *j* occurs after *i*. We consider lags up to five weeks. The null hypothesis is that the epidemic for region *i* does not Granger cause the epidemic in region *j*.

### 2.6 Nowcasting regionalization

To evaluate the potential for the prediction of regionalization within seasons, we applied our network-based regionalization approach on partial time series through the epidemic season. For every week of an epidemic which lasts for *T* weeks, *t ∈* [0, *T*], we inferred a network and performed robust community structure detection of county-level time series from weeks 0 to *t*. To evaluate nowcasting potential, we measure the region-specific recall (or sensitivity) of the region membership based on the complete time series by the region membership identified by the partial time series at every week, *t*. Recall is defined as the number of true positives divided by the sum of true positives and false negatives. Thus, region-specific recall is based on the number of counties correctly identified (based on a partial time series) to be part of a region divided by the number of counties truly in the region (as identified by the complete time series).

#### 2.6.1 Forecasting regionalization

To evaluate whether epidemic regionalization could be predicted across seasons, we used a machine learning approach. For each influenza season in our study period, we gathered county-level data on previously hypothesized drivers of influenza transmission. We considered 15 previously described county-level covariates [1] including vaccine coverage in toddlers and elderly, the distribution of A/H3 and B influenza subtypes, prior immunity, age distribution, household size, population density, poverty, healthcare access, insurance coverage, and healthcare-seeking rates. Additionally, we included covariates on the latitude of the county centroid, the Euclidean distance from most probable source county, and the pre-epidemic anomaly in specific humidity (See supplement for details).

We tested the ability for random forest models to predict clusters in future influenza seasons. For each influenza season, we trained a random forest model composed of 2000 trees to a cluster-stratified random sample of 85% of observations [35]. Model fit was evaluated by comparing classification error in the training set and 15% test set. For each influenza season model, we then predicted influenza season classifications for the following season (e.g., the 2006-2007 model was used to predict the 2007-2008 season clusters) and calculated prediction error as the percentage of county pairs that were incorrectly predicted to belong to different clusters.

We also sought to identify whether any covariates were consistently important classifiers of influenza season clusters. We measured variable importance with the mean decrease in accuracy, which represents the mean decrease in model classification accuracy after permuting the covariate [35]; larger values indicate covariates with greater potential importance to model prediction.

For sensitivity analysis, we compared the model performances for fit, prediction, and variable performance with an analogous set of models with a cluster-stratified random sample of 70% of observations.

## 3 Results

Our goal is to characterize regions with similar epidemic dynamics which can be treated as discrete geographical units in the implementation of public health policies. To achieve this, (a) we inferred U.S. county-level epidemic time series using medical claims data collected from 120,000 physicians totalling 2.5 billion visits for flu seasons during 2002-2008; (b) represented correlations in these time series as a complex network, and analyzed the structure of the inferred networks; (c) used a network theory approach to identify groups of epidemic time series (and thus locations) that are more dynamically correlated within than between; and (d) considered predictive models on this regionalization (Figure 1). Here, we present our results on the performance of our method, the structure of the epidemiological dynamics, and regionalization patterns across scales.

### 3.1 Spatial Structure of Epidemiological Dynamics

We analyze the inferred spatial network structure in which nodes represent U.S. counties and an edge between a pair of nodes represents that the epidemic time series in the two counties are similar with a Pearson’s correlation coefficient of 0.8 or above (Figure 2A) shows the network visualization for the season 2005-2006 (visualizations for other seasons can be found in the supplement).

**Figure 2:**
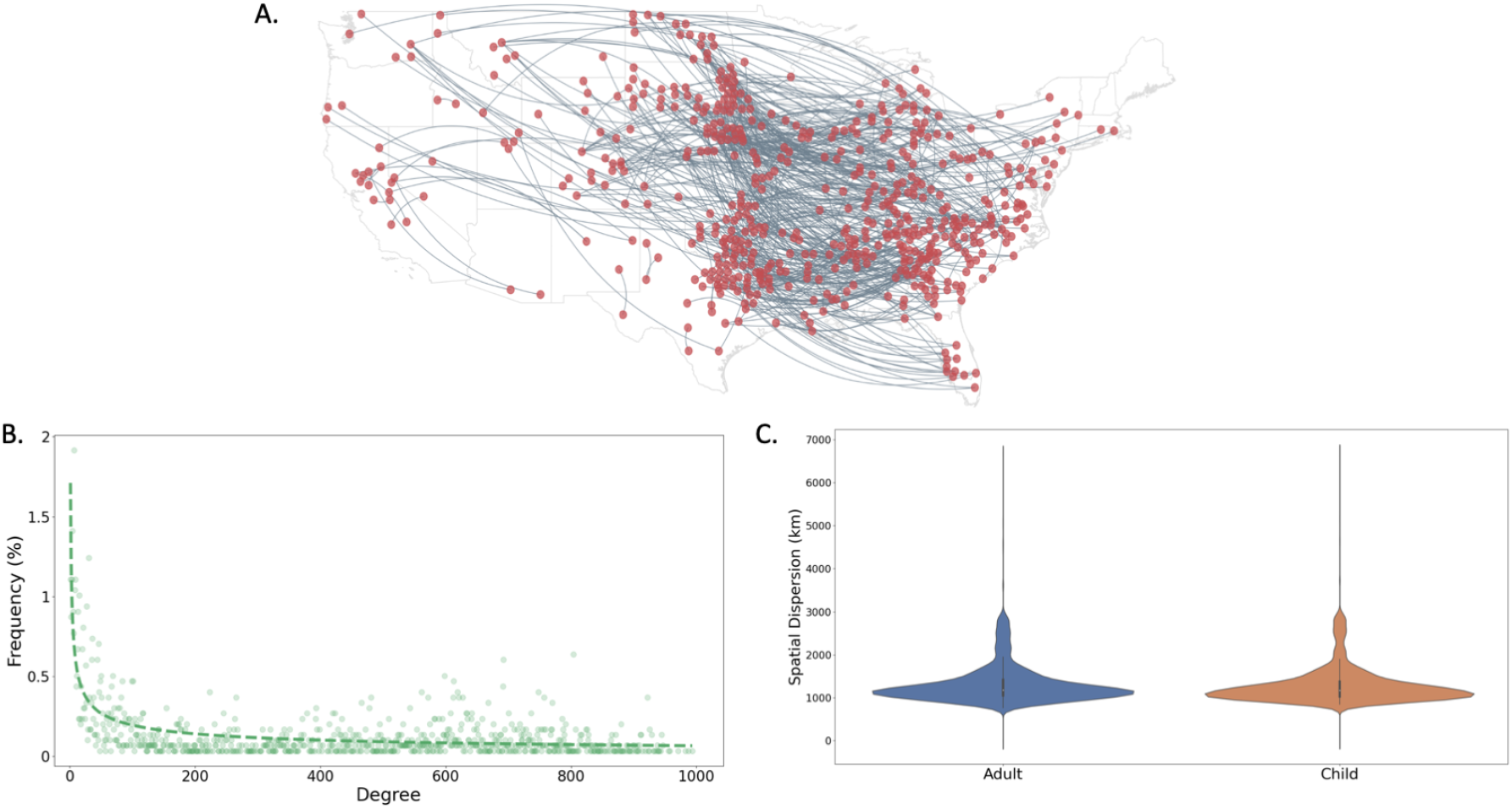
Spatial structure of the epidemiological network. (A) We visualize a sample of the epidemiological network for the 2005-2006 influenza season in which nodes are US counties and edges represent epidemic time series similarity. (25% of western counties and 1% of eastern counties are shown). (B) The probability distribution of degree (i.e. node connectivity) for the 2005-2006 influenza season epidemiological network is highly skewed. (C) The spatial dispersion of edges in the 2005-2006 epidemiological network do not show variation by demographic group.

We show the degree distribution (Figure 2B for the 2005-2006 season (degree distribution plots for other seasons can be found in the supplement). All seasons show power law behavior meaning that most counties have low degree and share epidemic dynamics with only a few other counties, while a few counties have very high degree and have similar epidemic dynamics to a large number of locations. We also find that the mean degree varies greatly by season (e.g. 170 in 2005-2006 and ∼ 720 in 2007-2008), suggesting that the heterogeneity of epidemic dynamics is larger in some seasons than others.

We separately compare the the spatial network inferred for the adult population to that of the child population. Our hypothesis was that the adult population network would be more spatially dispersed than the child network. We find that the distribution of spatial dispersion across network edges has a distribution centered on connectivity at 1000 kilometers (∼ 600 miles) (Figure 2C), and that there is no statistically significant difference in spatial dispersion between the two demographic groups.

### 3.2 Regionalization of Epidemiological Dynamics

We constructed robust epidemiological geographies based on epidemic incidence data from influenza seasons during 2002-2008. To measure the performance of our regionalization results, we calculated the modularity of the epidemic similarity network. Relative modularity is high (ranging from 0.39-0.79 with an average of 0.59) across seasons, as are the silhouette scores (details can be found in Figure S1). We also compared the regionalization results from our network-based method to those from the popular method of hierarchical agglomerative linkage clustering. We find qualitatively similar results with an average normalized mutual information score of 0.43 (Please see Table S3 in the supplement).

The results of our network-based community structure detection identify a regionalization of U.S. counties where each region experiences similar epidemic dynamics. The method results in a structurally optimal regionalization which may not be feasible for public health implementation. We thus spatially smooth the regionalization post-hoc to achieve more results that would be more feasible for public health implementation given their geographic cohesiveness, while losing some network quality. Figure 3A and 3B shows the results for the 2005-2006 season before and after the smoothing for feasibility (maps for other seasons can be found in Figure S4 in the supplement. We note that the feasibility adjustment does not result in significantly different partitions (with normalized mutual information scores of 0.77-0.87 (see Table S6).

**Figure 3:**
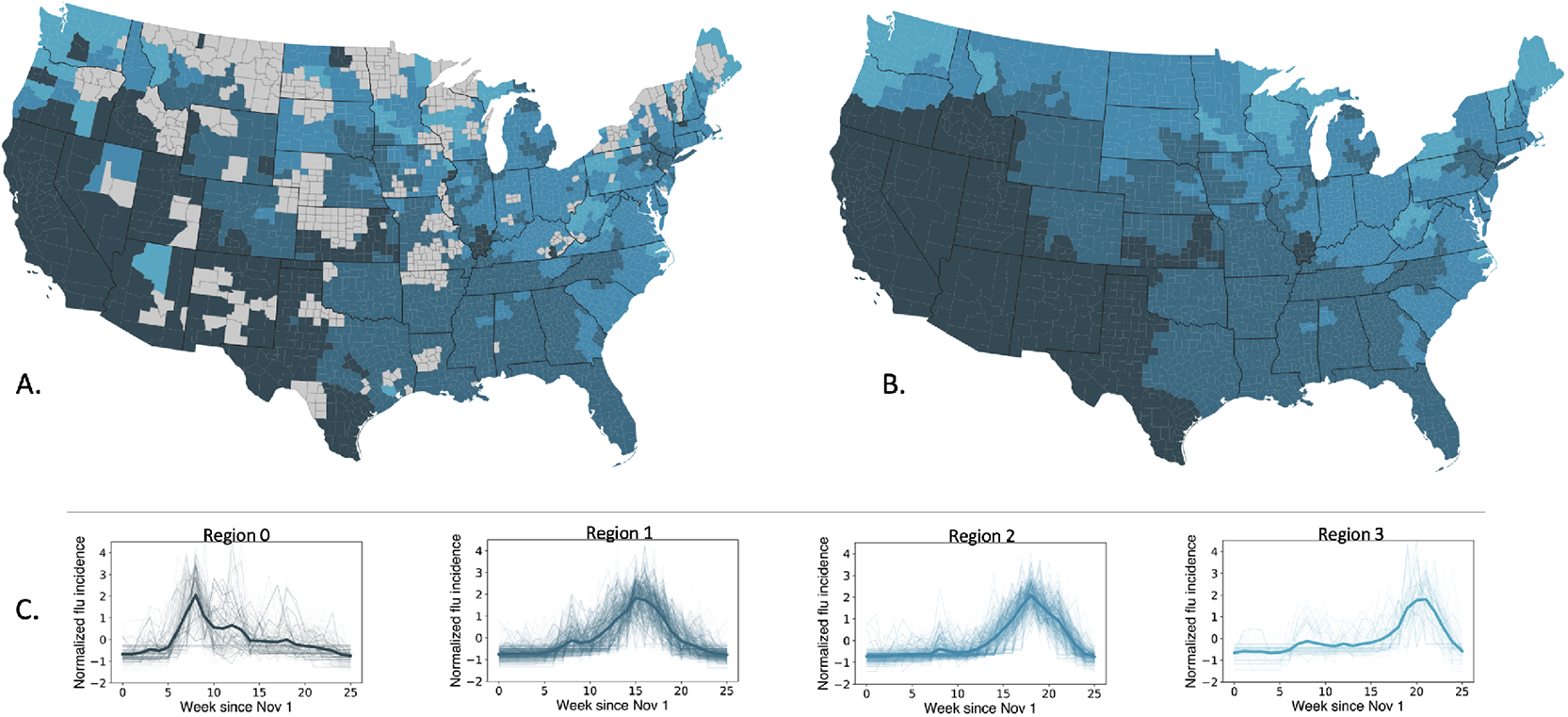
A) The robust community structure partition identified by the Louvain algorithm and our robustness analysis for the 2005-2006 influenza season. B) The epidemiological regionalization for the 2005-2006 influenza season generated by making the theoretically optimal community structure shown in panel A more feasible for public health implementation. Counties of the same color belong to the same epidemiological region. C) The epidemic time series of all US counties during the 2005-2006 influenza season grouped by region based on the regionalization shown in panel B. Color shades denote the timing of epidemic dynamics with lighter shades denoting successively later epidemic dynamics.

We consider the epidemic dynamics of each region separately (Figure 3C), and find that the regionalization corresponds to different waves of influenza outbreaks during the season. We find that the time series of the epidemics in each region are epidemiologically distinct from others, and regions experience statistically different onset and peak times separated by one or more weeks (t-test results can be found in Table S2. We find that the regions are also highly cohesive with an average intraregion connectivity of 70% and an average intraregion similarity of 88% across regions and seasons. (Full results can be found in Figure S4). The identity of the regions thus captures the progression of the epidemic through the country, making this of epidemiological value.

We hypothesized that the regionalization patterns across influenza seasons would be consistent, as past work has shown that influenza epidemics start in the southern United States and progress northward[4]. Our results show that the progression of the influenza season across the country in fact varies significantly across seasons (Figure 4). We find that the earliest region in the country to experience influenza is often in the southwest (2005-2006, 2007-2008, 2008-2009 seasons) but can also be in the southeast (2006-2007 seasons) or in the central south (2002-2003 season). Epidemics thus progress from west to east in a one-dimensional wave (as in the case of a southwest start) or have more complex progressions. We also find that seasons fall into three classes with respect to the respective timing of epidemics across regions: in the 03-04 season, all regions peak early; in the 04-05, 07-08, and 08-09 seasons all regions peak late; and during the 02-03, 05-06, and 06-07 seasons, regions are distributed in their peak timing (Table S1). We also hypothesized that regionalization patterns would vary by demographic group. However, we do not find any qualitative differences in the regionalization patterns.

**Figure 4:**
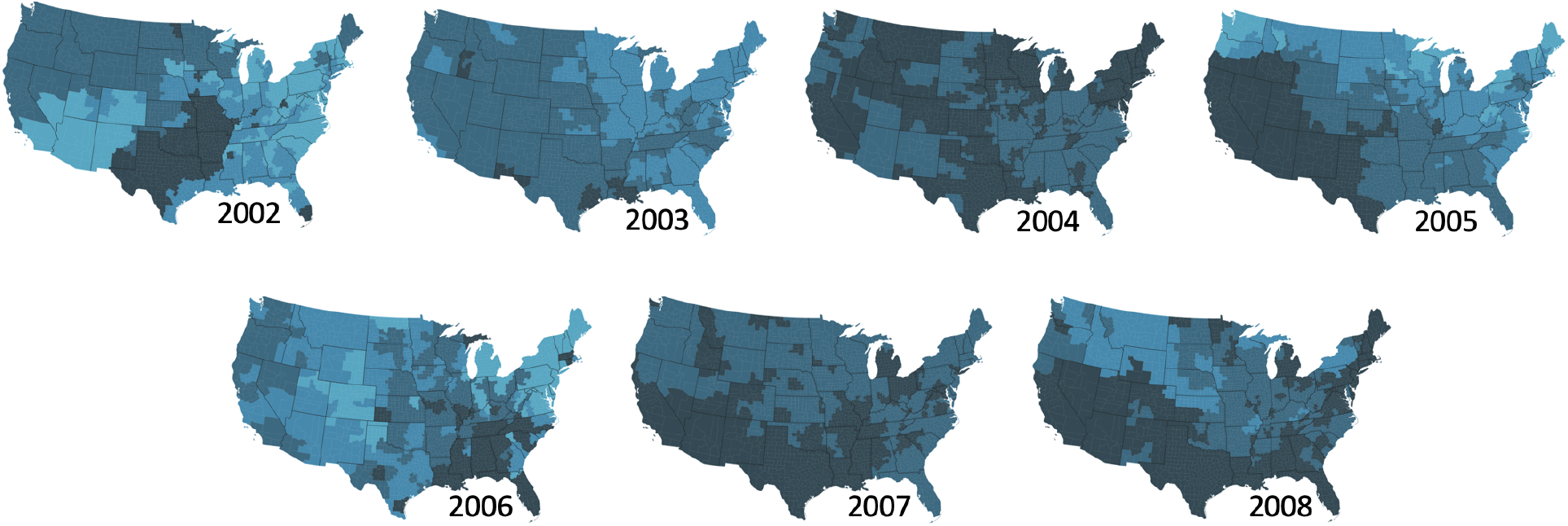
The epidemiological geographies for influenza seasons from 2002 to 2008. The regionalization shown is the result after our community structure detection, robustness analysis, and feasibility analysis. Colors represent the relative timing of the epidemic for each community with darker colors representing earlier epidemic dynamics.

Lastly, we compare our results to the existing regionalization used by the U.S. Department of Health and Human Services (HHS) for resource allocation and public health administration. We find that our epidemiologically-driven regionalization does not show much consistency with the administrative regionalization of the HHS regions (Figure S7 and Table S7).

### 3.3 Prediction of Regionalization

Our results suggest that our method of epidemiological regionalization produces regions that correspond to the progression of the influenza epidemic through the country. We consider three follow-up questions based on this result: a) Can epidemic dynamics from one region be used to predict the epidemic dynamics of the next region?; b) Can regionalization patterns be nowcast through the course of an epidemic? and c) How can we predict regionalization patterns across seasons based on demographic, social and environmental processes? The answer to the first question can help us understand the epidemic forecasting potential of the region-specific epidemic time series; and the answer to the second and third question can allow for the prediction of regions based on partial epidemic data or covariate data.

To address the forecasting potential for epidemic dynamics, we use the Granger causality test. We find that regions with earlier outbreaks do indeed Granger cause the outbreaks that occur subsequently in other regions. That is, past values of an earlier region’s epidemic time series have a statistically significant effect on the current value of a later region’s epidemic, taking past values of the later region’s outbreak into account. However, we did not find that this was true for all lags, making this a less robust result. (Details can be found in S8).

To evaluate the potential to nowcast regionalization within an influenza season, we applied our network-based regionalization approach to partial epidemic data. We find that while partial epidemic data is not able to identify all regions given the lags in epidemic trajectories between regions (Figure 5A). However, the partial regionalization succeeds in characterizing individual regions, and this success is correlated with the timing of the epidemic in each region– the membership of each region is resolved as the region reaches its epidemic peak (Figure 5B). Thus while epidemic data from a full season is necessary to characterize a national regionalization, the identification of individual regions earlier in the season provides information about the regions of the country which will face later epidemics.

**Figure 5:**
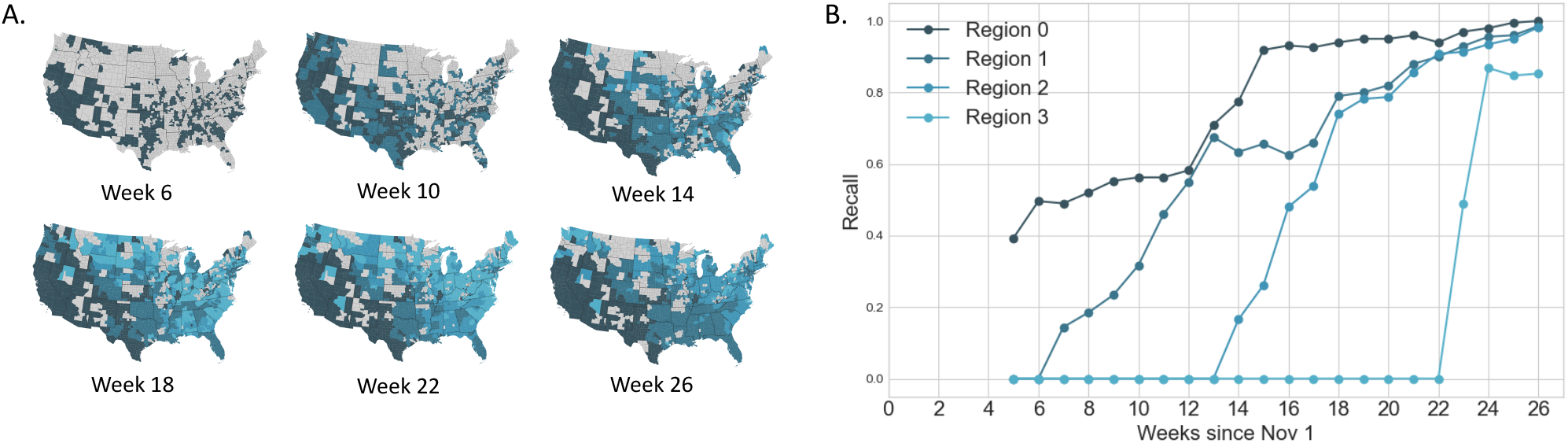
A) We consider regionalization results based on partial epidemic time series data up to Week 6, 10, 14, 18, 22, and 26 starting at November 1st for the 2005-2005 influenza season. B) For every week, *t*, we consider the region-specific recall (or true positive rate) for a regionalization based on partial epidemic time series with data from week 0 to *t*. The membership of each region is resolved as it reaches its epidemic peak.

Random forest models were used to quantify the predictability of future regionalization clusters for previous ones. Models were first fit to clusters for an individual season, and model performance was assessed by comparing test error to out-of bag error for the regionalization classifications of each influenza season model (Table S8). Both test and out-of-bag errors ranged from 8 to 14% across models. Models were then used to predict the regionalization clusters for one-season-ahead, and the prediction errors across models ranged from 28% (2006 model predicting 2007 regionalization clusters) to 61% (2002 predicting 2003) (Table S9). Variable importance was quite consistent across influenza seasons, with distance to most probable source location, latitude, elderly vaccination coverage, and proportion of H3 circulation contributing to the greatest mean decreases in accuracy with relative stability across models (Figure 6). Results were consistent in sensitivity analysis (See supplement for details).

**Figure 6:**
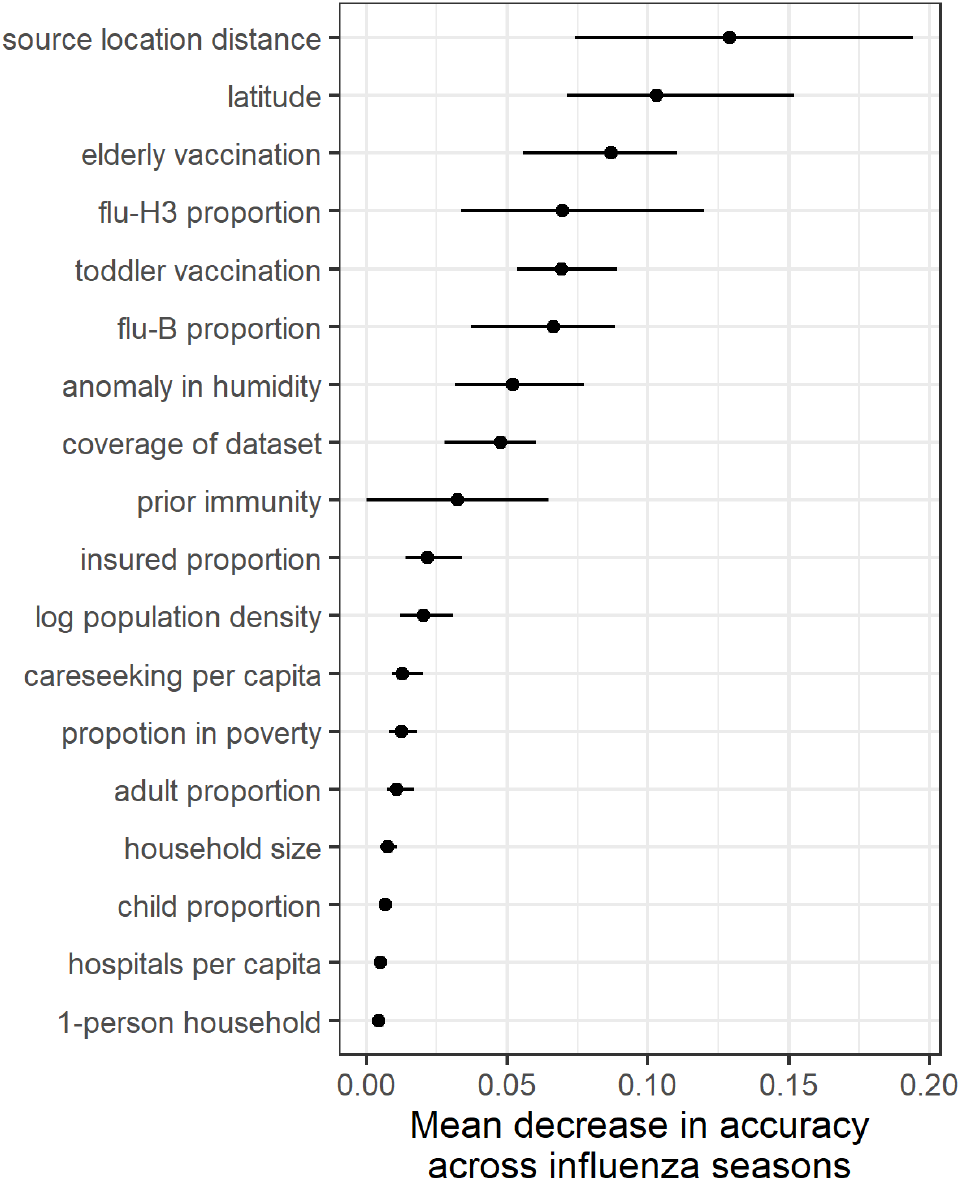
Variable importance for random forest classification of influenza season clusters. Variable importance is represented as the mean decrease in accuracy across influenza seasons (point is mean of the mean decrease across seasons, line range is the range of mean values across seasons).

## 4 Discussion

Infectious diseases like seasonal influenza exhibit highly dynamic and complex spatio-temporal patterns, and our understanding of these dynamics remains limited. Current public health surveillance and mitigation efforts rely on the assumption that infection diseases respect administrative or geopolitical borders, when they have been shown to not (e.g. [36, 37, 38, 39]). In this study, we have presented a quantitative method of identifying a fine-scale regionalization that is based on epidemiological characteristics rather than geopolitical boundaries. Our results suggest that seasonal influenza regionalizes the United States into three to five epidemiologically distinct regions. All locations within each spatial region experience synchronous influenza epidemics, and the outbreak exhibits wave-like propagation with an average ∼ 2 week delay between regions. However, we find that there is significant heterogeneity across flu seasons in the identity of the regions and the relative timing of epidemics across regions and that the regions from one season are not predictive of those in future seasons. Our work is also consistent with the recent work of Polyakov et al. who used a similar methodology to explore spatial optimization of an epidemic alert system in France [40].

This regionalization approach necessitates a high-resolution source of epidemiological data that can distinguish transmission dynamics at smaller spatial scales. The U.S. ILI outpatient ILI surveillance system (ILINet) is composed of roughly 3,000 sentinel physicians in the United States. While this surveillance system may be sufficient for capturing the broad geographical distribution of ILI during the influenza season, these data may not provide the geographic resolution necessary to infer geographical clusters that are useful for public health practice. Non-traditional sources of ILI data such as medical claims provide a high volume and high-resolution alternative to ILI sentinel surveillance, and previous work has validated the similarities between the ILI patterns observed in medical claims data [13] and shown its utility in re-thinking ILI surveillance design [1].

Our time series clustering approach relies on defining time series similarity with the Pearson correlation coefficient [41]. Shape-based clustering approaches, such as ours, are expected to be more effective compared to feature-based approaches as they incorporate information about the entire time series, and can be used across different times and scales [42]. Our interest in the synchronicity of epidemic outbreaks made the unit-independent Pearson correlation the appropriate measure. Our approach is, however, generalizable to other time series distance measures if other aspects of the dynamics are of interest [41]. We interpret the time series similarities through a complex network representation and apply a well-established community detection algorithm. This clustering method has been shown to outperform traditional time series clustering algorithms [43].

The communities in a disease dynamics-driven regionalization may not correspond to administrative boundaries nor be spatially contiguous areas, and are in fact the result of a variety of complex factors such as environmental variability and human mobility. Recent efforts have shed light on the socio-environmental factors that may drive spatio-temporal seasonal influenza dynamics in temperate zones [1, 2, 3, 4], but a thorough understanding remains elusive. The advantage of our method is that it identifies communities without knowledge of these factors or a mechanistic understanding of them, and is instead based simply on empirical disease dynamics. In the absence of such an *a priori* understanding, our method can be used to make some inferences about mechanism. Indeed, the wave-like propagation and relative spatial cohesiveness of the epidemiological regionalization produced by our method suggests that counties proximal to each other often experience similar influenza epidemics, suggesting that geographic proximity is an important driver for influenza dynamics. The consistent importance of the distance from source location and latitude in explaining the regionalization clusters further corroborates previous studies that have characterized the hierarchical dynamics of seasonal influenza [14, 15, 2] in which transmission decays as geographic distance between two locations increases [2] and transmission is mainly driven by local commuting and work flow patterns rather than long-distance or airline mobility [2, 5, 15, 4]. Surprisingly, our regionalization results did not provide any evidence for our mobility-based hypothesis for more dispersed spatial dynamics of ILI among adults.

Our work also confirms that most influenza epidemics start in the southern U.S. [2, 4], and some seasons (e.g. 2005) match the west to east pattern proposed by Wenger et al. for influenza (among adults over 65) [44]. Additionally, the regionalization reveals a unique feature of epidemic timing across regions in which seasons are homogeneously early or late, or heterogeneous in their peak timing across regions. We note that this partitioning of influenza is consistent with the severity classification we proposed in [45] and suggests that severe seasons are homogeneously early, moderately severe seasons are uniformly late, and mild seasons are distributed in their timing, which is consistent with a previous study by Dahlgren et al. [46]. The wave-like progression of flu through the U.S. also allows for the prediction of epidemic dynamics in a region based on other regions (as demonstrated by our Granger causality results) and for the prediction of region membership during the course of the epidemic (as highlighted by our partial regionalization results). This finding shows that it is possible to provide an understanding of how regions are shaping over time, yielding expected trajectories of the epidemic evolution in different parts of the country. Resources can potentially be mobilized according to the accruing of information provided by the method, together with communication targeting areas with soon-to-be-expected peak or those experiencing a delayed epidemic. This information cannot be easily gathered by simply looking at county-level surveillance data, because of the intrinsic noise and possible changes of tendency. A higher level analytical tool that is able to have a systemic view to identify similarities across space is therefore needed to identify the robust signal beyond the local noise and guide planning in real-time.

The epidemiological dynamics of influenza have long been modeled with metapopulation models [47, 48, 49]. Such models rely on the definition of subpopulations which are assumed to exhibit homogeneous disease dynamics within. The optimal spatial scale of these subpopulations, however, remains unknown. This spatial scale relates to the *characteristic scale* for influenza dynamics, and can be thought of as the spatial resolution at which maximum spatial variance exists between locations and minimum variance exists within locations. The question, for the US for example, is: are influenza dynamics homogeneous on the scale of counties, states, or multiple states? We argue that our regionalization approach provides a means to address this question and suggests that the characteristic scale of influenza is quite large and spans multiple states, albeit not following administrative boundaries. (However, we note that since our data are limited to a spatial resolution of U.S. counties, we may not be able to identify heterogeneity that may exist at finer scales [50].) In countries and regions smaller than the U.S., influenza dynamics have been shown to be synchronous nationally and internationally [51, 52, 40], also suggesting a characteristic speed for disease movement. Future work on better understanding the spatial scales of transmission can inform the scales of intervention strategies to prevent local intensification and regional propagation of disease [53].

Some data and methodological limitations affect our study. First, we use ILI data as a proxy to understand the spatio-temporal dynamics of influenza, but this has been known to be a reliable method for characterizing influenza incidence [14]. Second, our data are derived from healthcare interactions and thus may be subject to spatially-varying bias. We have mitigated this concern by normalizing our data on ILI visits by metadata on healthcare visits for any cause. This normalization helps capture spatial and temporal variation in healthcare access, insurance coverage and healthcare-seeking behavior. We also note that while these biases may affect time series amplitude significantly, the period (which is of relevance to our study) is less likely to be affected. Third, our regionalization results depend on the edge weight (i.e. time series correlation) threshold chosen. However, recent work demonstrates that community structure is preserved with edge weight thresholding [54], and we highlight that the regionalization results across thresholds are related hierarchically (Figure S2).

Our method of using fine-scale epidemiological data to inform regionalization has the potential to inform disease prevention and management for ILI. We compare our regionalization to the current regions used by HHS for public health management, and find that epidemiological dynamics within HHS regions are highly heterogeneous, suggesting that homogeneous policy is unlikely to be efficient (Figure S7). Our approach more accurately groups areas that have similar epidemic epidemics, using a systemic approach while also identifying a simple heuristic (peak time) that differentiates epidemic regions which may be more readily used in public health contexts. While our work highlights that our ability to predict influenza regionalization across seasons remains limited (perhaps due to limited data), there is potential to use our approach to inform real-time regionalization during a season to divert resources to regions yet to experience outbreaks. We find that influenza regionalization may be partially explained by epidemiological factors such as elderly vaccination coverage, suggesting it may be beneficial to group these locations for vaccination prioritization. Our results also suggest that planning the timing of vaccine distribution based on regionalization is likely to minimize the effects of waning immunity [55]. Furthermore, optimal regionalization results can also inform novel surveillance approaches [56], as well as to plan efficient coordination between local health departments [57]. Public health preparedness and response in the face of high heterogeneity demands effective but tractable approaches; regionalization as a strategy has the potential to balance both.

## Data Availability

All data associated with this study are provided in a GitHub repository:

https://github.com/bansallab/fluregionalization

## Acknowledgments

This work was supported by the Jayne Koskinas and Ted Giovanis Foundation for Health and Policy; the Research and Policy for Infectious Disease Dynamics (RAPIDD) program of the Science and Technology Directorate, Department of Homeland Security (DHS), and the Fogarty International Center, National Institutes of Health (NIH);and by the National Institute of General Medical Sciences of the NIH under Award Number R01GM123007. The content is solely the responsibility of the authors and does not necessarily represent the official views of the National Institutes of Health.

## SUPPLEMENT

### 1 Data Source

Weekly visits for influenza-like illness (ILI) and any diagnosis from October 2002 to April 2009 were obtained from a records-level database of US medical claims managed by SDI Health (later known as IMS Health and now known as IQVIA) and aggregated to three-digit patient U.S. zipcode prefixes (zip3s), where ILI was defined with International Classification of Diseases, Ninth Revision (ICD-9) codes for: direct mention of influenza, fever combined with respiratory symptoms or febrile viral illness, or prescription of oseltamivir. Medical claims have been demonstrated to capture respiratory infections accurately and in near real-time [58, 59], and our specific dataset was validated to independent ILI surveillance data at multiple spatial scales and age groups and captures spatial dynamics of influenza spread in seasonal and pandemic scenarios [13, 1].

We also obtained database data from SDI Health on total visit volume for any cause (“all cause visits”). ILI visits and all cause visits were redistributed to the county-level according to population weights derived from the 2010 U.S. Census ZIP Code Tabulation Area (ZCTA) to county relationship file, assuming that ZCTAs that shared the first three digits belonged to the same zip3 [1]. We normalize the time series of ILI visits by all cause visits to account for spatio-temporal variation in healthcare access, healthcare seeking, and database coverage. Epidemic time series are defined for the period of November 1 to April 30. For the 2008-2009 flu season, we restricted our data to March 31 given the late dynamics of the H1N1 pandemic. To summarize the epidemic characterisitcs for validation, we defined onset and peak times. The epidemic period was the consecutive period bookended by periods where detrended ILI exceeded the epidemic threshold during the flu period wording [1]. We defined onset timing as the number of weeks from November 1 until the first week in the epidemic period. We defined peak timing as the number of weeks from week 40 until the week with the maximum epidemic intensity during the epidemic period.

#### 1.1 Data Ethics

Patient records and information in the medical claims dataset were anonymized, deidentified, and aggregated by SDI Health. All analyses were performed with aggregated time series data for influenza-like illness rather than patient-level information. Ethical review was sought from the Institutional Review Board at Georgetown University and was deemed exempt.

### 2 From Time Series to Complex Network

#### 2.1 Determining Similarity Threshold

When making the epidemic time series correlation networks, we chose to limit the network edges to time series correlations of 0.8 or higher. We chose this threshold to reduce the size and complexity of the network, to ensure that the network only included strong correlations, and because this threshold balanced high network statistics with epidemiological value. We evaluated a number of network metrics when determining the threshold including: silhouette score, modularity, percent coverage of all U.S. counties, and number of communities. The first two metrics measure how clustered the regions are, the latter two are to measure public health implementation effectiveness and ease. In Figure S1, we summarize modularity and silhouette values for each threshold for each year along with the percent of all U.S. counties in the resulting regionalization after any communities smaller than 94 counties (3% of all US counties) were removed, the number of communities for each threshold, and whether or not communities peaked during the same week. We identified the threshold of 0.8 by maximizing silhouette score, modularity, and coverage and minimizing the number of communities. Our chosen threshold of 0.8 also is appropriate for differentiating the regions epidemiologically in terms of peak timing (Figure S1). In Figure S2, we also illustrate that the regionalization across thresholds is quite similar.

**Figure S1:**
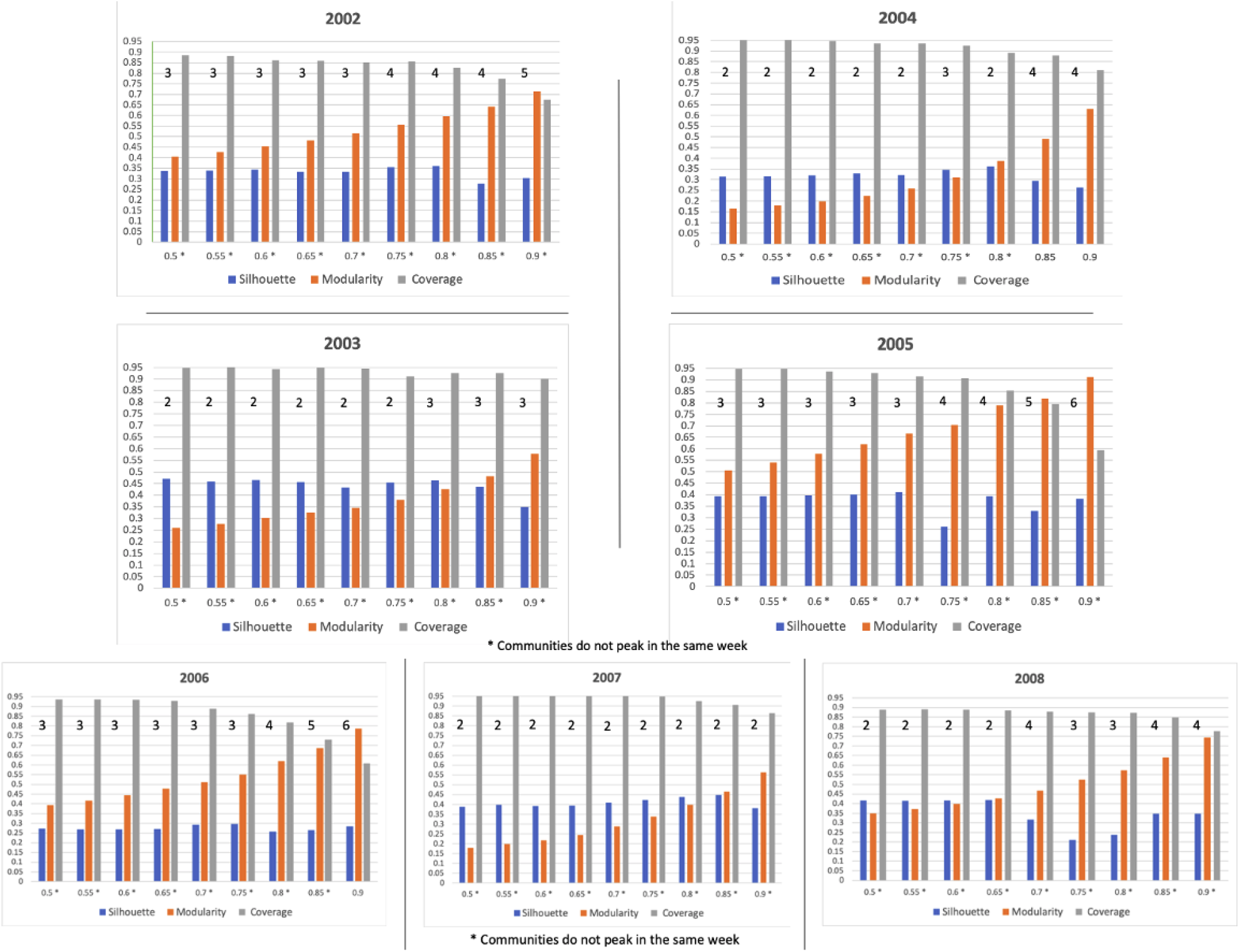
Determining Threshold for Network Structure. Figures showing the silhouette score, modularity, percentage of counties included in the final regionalizations, number of communities in each regionalization, and whether or not communities peak in the same week for each flu season in the study. A threshold of .8 was identified based on maximizing silhouette and modularity values, keeping coverage above 80% of U.S. counties. We disqualified thresholds of 0.85 and 0.9 based on having multiple years with coverage of less than 80%. Lower thresholds were disqualified based on their low network metrics and the fact that they did not fully highlight the granularity of the differences in the communities based on their low number of communities. Additionally, we highlight that the threshold of 0.8 also differentiates regions by peak timing.

**Figure S2:**
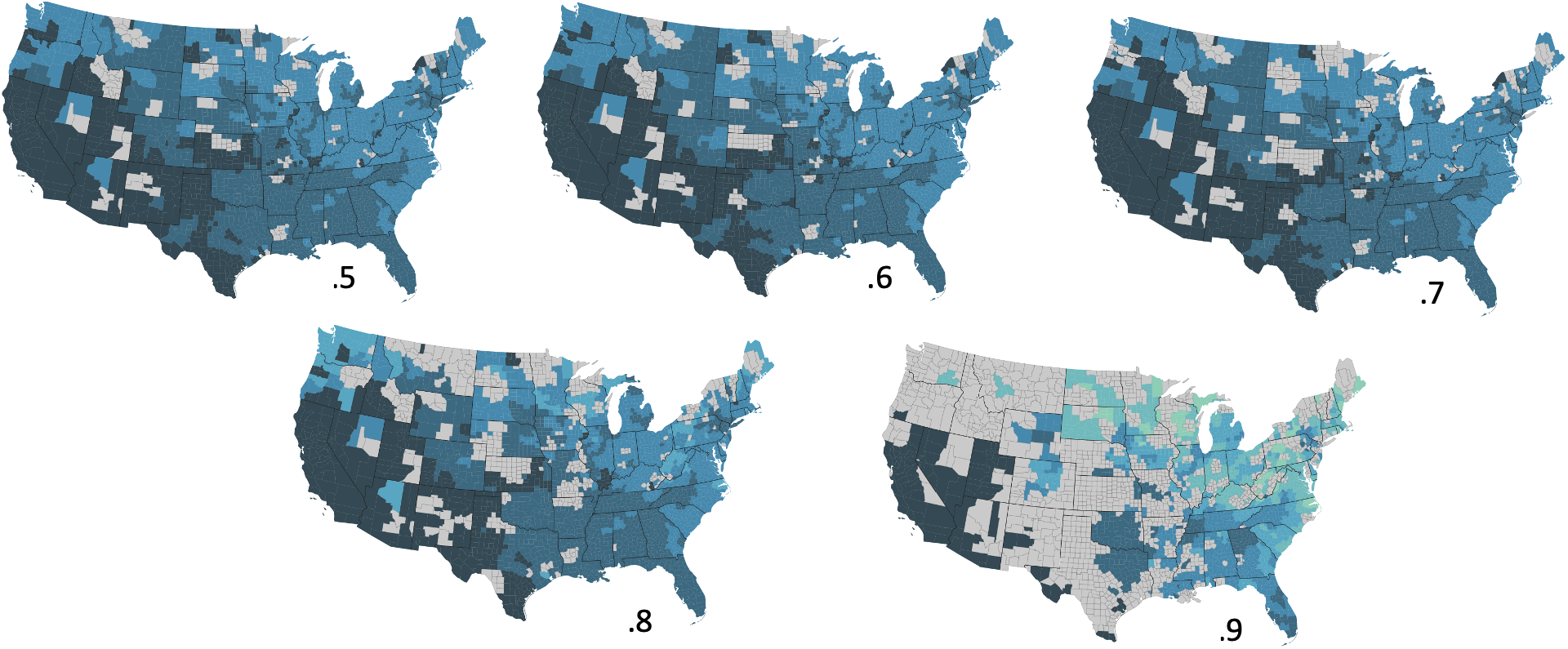
Epidemiological regionalization for the 2005-2006 flu season using different thresholds for the time series similarity network (e.g. a threshold of 0.5 means that only pairs of counties with a Pearson’s correlation ≥ 0.5 are connected in the similarity network). The regionalization across thresholds is consistent.

### 3 Validating Community Structure

We validated our method epidemiologically and methodologically. Our epidemiological validation consisted of analyzing the timing of two important epidemiological characteristics, the onset time and the peak time, as an average for each community. Please see Table S1 for a list of average peak weeks and onset weeks for each community in each year using a threshold of 0.8. Furthermore, we evaluated whether or not the peak times were statistically different across communities for a flu season by running a two-sided t-test, please see Table S2.

Our second method of validation compared our results based on the Louvain algorithm (our chosen algorithm) with another common clustering method to test for sensitivity to the method. We used average linkage hierarchical clustering with Pearson’s correlation as the distance function. Figure S3 shows a disease geography for the 2005-2006 flu season for both the Louvain algorithm and the hierarchical clustering algorithm showing very similar but not identical geographies. Furthermore, we quantified this variation by calculating the mutual information between the two regionalizations (Table S3), and find that the two methods generally find consistent regionalization results.

### 4 Evaluating Community Structure

Following the identification of the community structure in the epidemic time series similarity networks, we evaluated the structure of the communities, the mean correlation of epidemic time series within each community or region, the standard deviation of those correlations, and the within-degree ratio of the regions. Table S4 shows that the correlation of time series within each community was high, ranging from 86-90% which suggests that the community structure characterized regions exhibiting highly uniform epidemics. Additionally, we found that the regions were highly cohesive with within-degree ratios of 54-98% which measures the ratio of degrees within a given community compared to those outside of the community.

**Table S1:** Comparing Average Onset and Peak Time. Here is a table of the average epidemic onset time and peak time for each community in every flu season using the .8 threshold results. No flu season has communities that have onset or peak times in the same week suggesting epidemiologically-significant differences in dynamics between communities.

| Average Onset | Average Peak | Community Number | Flu Season |
| --- | --- | --- | --- |
| 4.17 | 8.57 | 0.0 | 2002.0 |
| 7.00 | 12.74 | 1.0 | 2002.0 |
| 8.36 | 14.28 | 2.0 | 2002.0 |
| 11.44 | 17.00 | 3.0 | 2002.0 |
| 0.0 | 0.33 | 0.0 | 2003.0 |
| 0.97 | 4.74 | 1.0 | 2003.0 |
| 2.57 | 6.82 | 2.0 | 2003.0 |
| 6.44 | 14.24 | 0.0 | 2004.0 |
| 7.45 | 16.04 | 1.0 | 2004.0 |
| 4.59 | 9.32 | 0.0 | 2005.0 |
| 6.99 | 15.22 | 1.0 | 2005.0 |
| 10.09 | 18.19 | 2.0 | 2005.0 |
| 11.78 | 20.27 | 3.0 | 2005.0 |
| 2.77 | 10.48 | 0.0 | 2006.0 |
| 6.54 | 14.07 | 1.0 | 2006.0 |
| 8.25 | 15.61 | 2.0 | 2006.0 |
| 8.72 | 17.62 | 3.0 | 2006.0 |
| 6.16 | 14.35 | 0.0 | 2007.0 |
| 7.53 | 16.52 | 1.0 | 2007.0 |
| 9.16 | 15.17 | 0.0 | 2008.0 |
| 12.19 | 17.15 | 1.0 | 2008.0 |
| 15.87 | 18.96 | 2.0 | 2008.0 |

**Table S2:** Evaluating Differences in Communities Based on Peak Time. Here we show the results of a Welch’s two-sided T test for comparing the differences in peak time between every community for every flu season. Communities are statistically different with p values less than .05.

| Community | Community | P Value | T Statistic | Flu Season |
| --- | --- | --- | --- | --- |
| 1.0 | 2.0 | 3.49E-227 | -38.36 | 2002.0 |
| 1.0 | 0.0 | 3.01E-162 | 39.83 | 2002.0 |
| 1.0 | 3.0 | 3.60E-196 | -55.71 | 2002.0 |
| 2.0 | 0.0 | 7.68E-227 | 53.25 | 2002.0 |
| 2.0 | 3.0 | 9.47E-132 | -34.04 | 2002.0 |
| 2.0 | 1.0 | 3.50E-322 | 48.07 | 2003.0 |
| 2.0 | 0.0 | 1.39E-123 | 117.13 | 2003.0 |
| 1.0 | 0.0 | 3.81E-157 | 65.92 | 2003.0 |
| 1.0 | 0.0 | 5.66E-238 | 36.65 | 2004.0 |
| 1.0 | 2.0 | 0.0 | -46.76 | 2005.0 |
| 1.0 | 0.0 | 2.41E-153 | 38.96 | 2005.0 |
| 1.0 | 3.0 | 8.05E-120 | -42.23 | 2005.0 |
| 2.0 | 0.0 | 5.31E-222 | 59.24 | 2005.0 |
| 2.0 | 3.0 | 6.21E-46 | -17.72 | 2005.0 |
| 0.0 | 3.0 | 6.04E-255 | -60.57 | 2005.0 |
| 1.0 | 2.0 | 2.55E-113 | -24.80 | 2006.0 |
| 1.0 | 0.0 | 9.14E-70 | 19.95 | 2006.0 |
| 1.0 | 3.0 | 1.17E-129 | -29.73 | 2006.0 |
| 2.0 | 0.0 | 1.99E-117 | 28.95 | 2006.0 |
| 2.0 | 3.0 | 5.52E-57 | -17.56 | 2006.0 |
| 0.0 | 3.0 | 9.01E-173 | -34.93 | 2006.0 |
| 1.0 | 0.0 | 0.0 | 49.34 | 2007.0 |
| 0.0 | 1.0 | 7.54E-271 | -41.08 | 2008.0 |
| 0.0 | 2.0 | 0.0 | -61.38 | 2008.0 |
| 1.0 | 2.0 | 1.47E-142 | -34.13 | 2008.0 |

**Table S3:** Evaluating similarity between the Louvain community detection algorithm and a hierarchical clustering algorithm. We show normalized mutual information scores between the Louvain partition and the hierarchical clustering partition for each flu season.

| Mutual Information | Flu Season |
| --- | --- |
| 0.59 | 2002.0 |
| 0.36 | 2003.0 |
| 0.07 | 2004.0 |
| 0.71 | 2005.0 |
| 0.53 | 2006.0 |
| 0.22 | 2007.0 |
| 0.48 | 2008.0 |

**Table S4:** Evaluating Regional Structure. We evaluated the structure of the regions comprising the epidemiological geographies for each flu season by measuring the mean correlation between time series within a region and the within-degree ratio of each region.

| Flu Season | Region | Mean Correlation | Std of Correlation | Within-Degree Ratio |
| --- | --- | --- | --- | --- |
| 2002 | 0 | 0.87 | 0.05 | 0.88 |
| 2002 | 1 | 0.88 | 0.04 | 0.76 |
| 2002 | 2 | 0.88 | 0.04 | 0.69 |
| 2002 | 3 | 0.86 | 0.05 | 0.65 |
| 2003 | 0 | 0.90 | 0.05 | 0.69 |
| 2003 | 1 | 0.89 | 0.05 | 0.61 |
| 2003 | 2 | 0.91 | 0.04 | 0.75 |
| 2004 | 0 | 0.88 | 0.04 | 0.63 |
| 2004 | 1 | 0.88 | 0.04 | 0.67 |
| 2005 | 0 | 0.88 | 0.06 | 0.98 |
| 2005 | 1 | 0.86 | 0.04 | 0.87 |
| 2005 | 2 | 0.87 | 0.04 | 0.87 |
| 2005 | 3 | 0.87 | 0.05 | 0.54 |
| 2006 | 0 | 0.86 | 0.04 | 0.62 |
| 2006 | 1 | 0.87 | 0.04 | 0.65 |
| 2006 | 2 | 0.88 | 0.04 | 0.58 |
| 2006 | 3 | 0.86 | 0.04 | 0.63 |
| 2007 | 0 | 0.89 | 0.05 | 0.62 |
| 2007 | 1 | 0.90 | 0.04 | 0.71 |
| 2008 | 0 | 0.88 | 0.04 | 0.74 |
| 2008 | 1 | 0.88 | 0.04 | 0.70 |
| 2008 | 2 | 0.89 | 0.05 | 0.59 |

**Figure S3:**
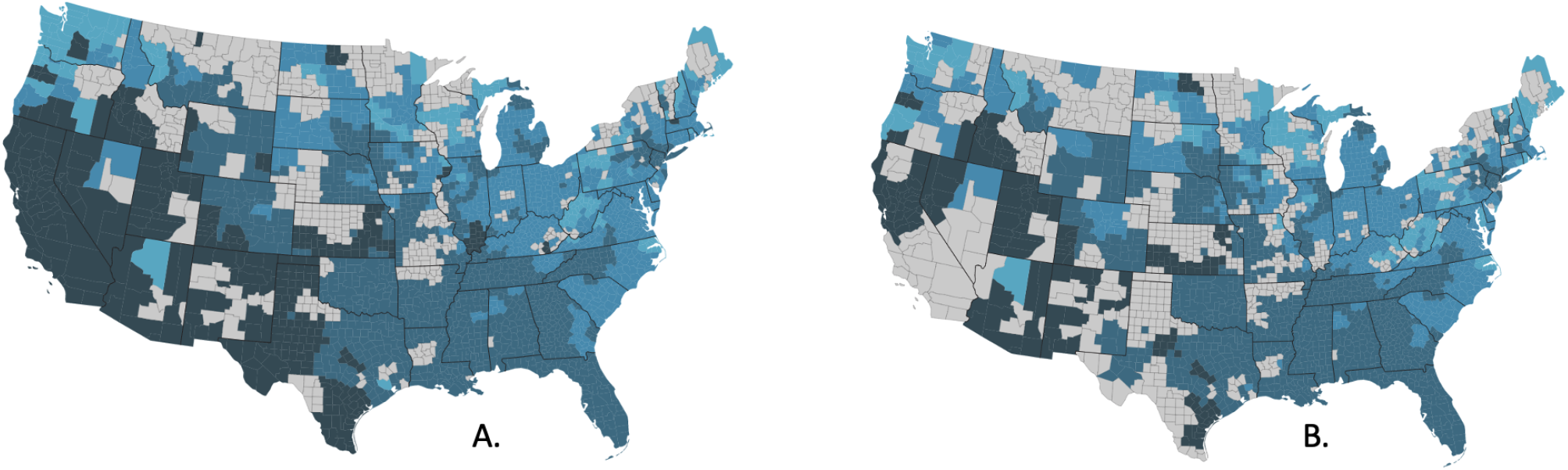
Validation of community structure by methodological comparison. We compared the resulting regionalization map for the 2005-2006 flu season from (A) the Louvain community structure detection (our method) to (B) an average linkage hierarchical clustering method.

**Figure S4:**
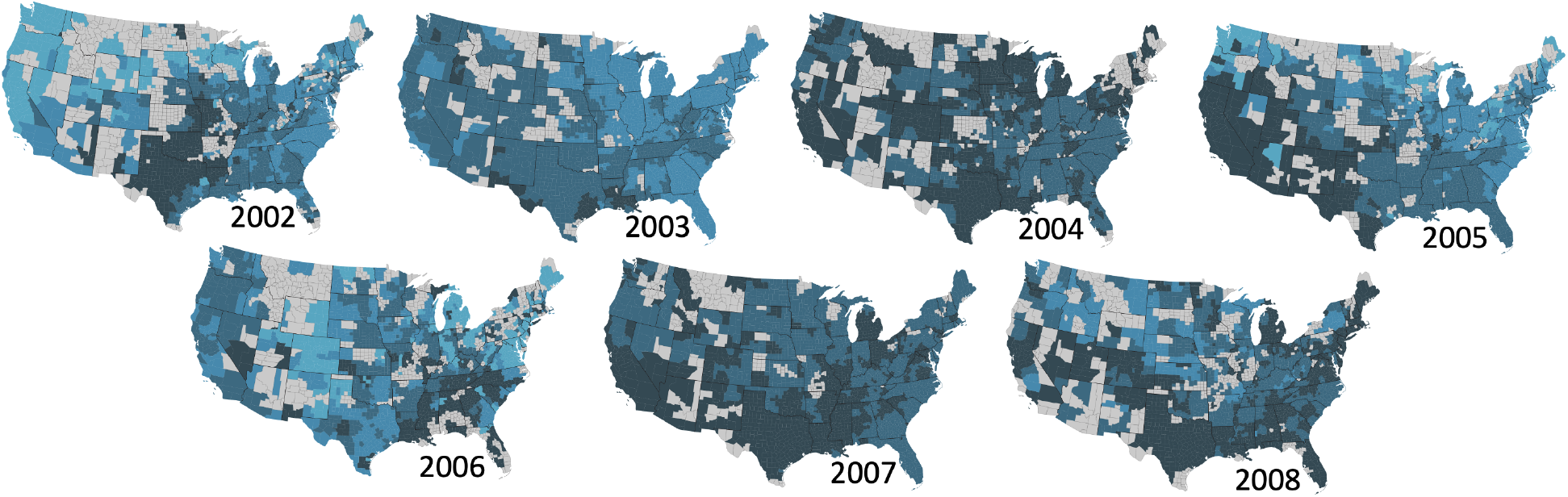
Louvain Regionalizations for all Years. Here we present all Louvain regionalizations for all flu seasons included in this study. These are the raw results before we applied our post-hoc feasibility analysis for the ease of public health implementation.

### 5 Louvain Regionalizations

We created regionalizations for each flu season (2002-2008) using the Louvain clustering algorithm. Please see Figure S4 for all maps for the 0.8 threshold.

#### 5.1 Evaluating Regionalizations

The disease geographies exhibit a high degree of heterogeneity across flu seasons. We quantified the variation in community structure by calculating the mutual information for every combination of flu season as seen in Table S5. Mutual information is low across seasons ranging from .005 to .1 suggesting a high degree of variation across years.

**Figure S5:**
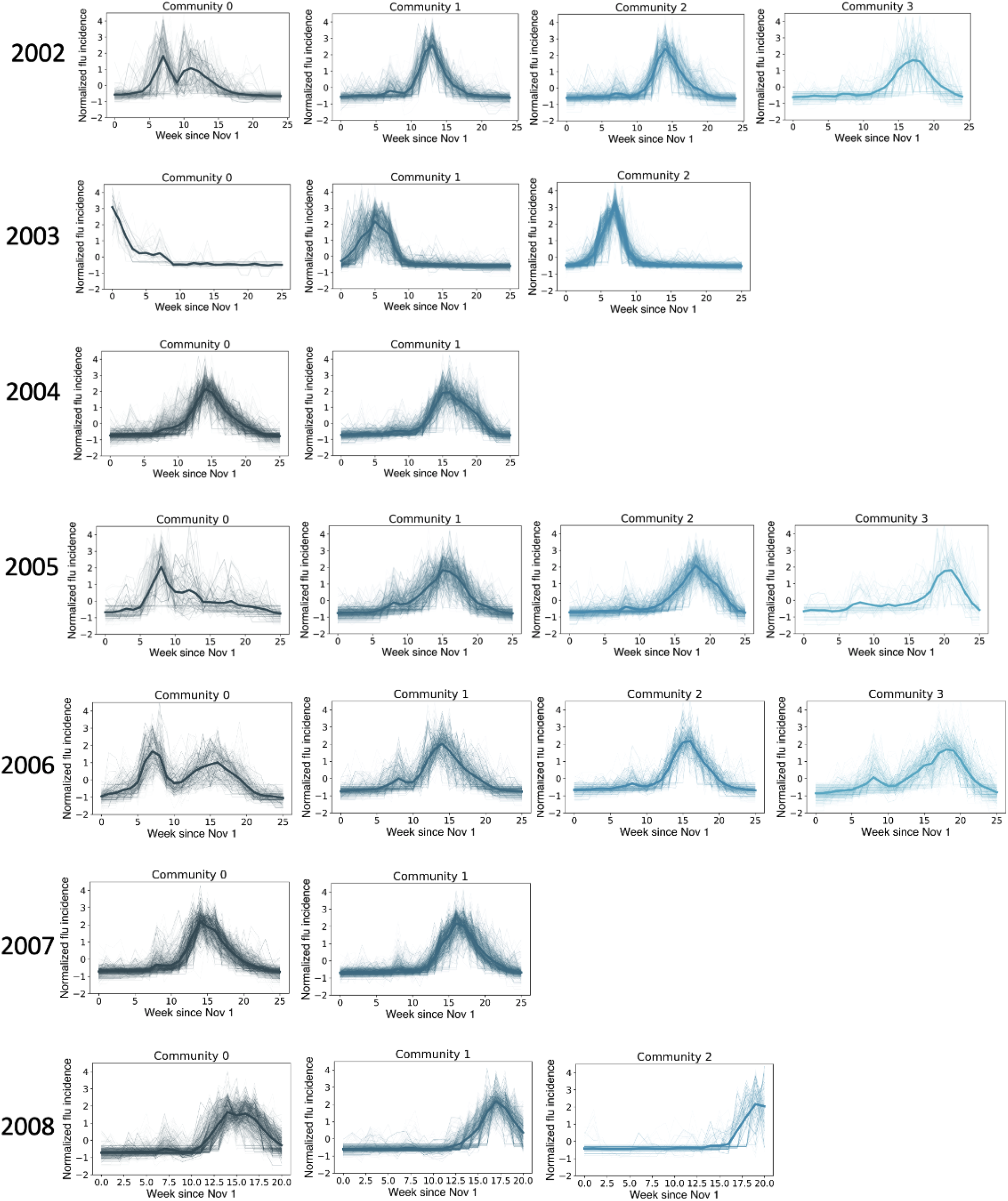
Time series for all communities in the 2002-2008 flu seasons.The average time series for each community is shown as a thick line.

**Table S5:** Comparing Normalized Mutual Information Scores between Seasons. Here we present the Normalized Mutual Information (NMI) scores between all combinations of flu seasons. NMI scores range from .005 to .1 which suggests that influenza seasons show significant heterogeneity across flu seasons and exhibit differential epidemiological characteristics from year to year.

| Mutual Information | Flu Season 1 | Flu Season 2 |
| --- | --- | --- |
| 0.079 | 2002.0 | 2003.0 |
| 0.024 | 2002.0 | 2004.0 |
| 0.101 | 2002.0 | 2005.0 |
| 0.069 | 2002.0 | 2006.0 |
| 0.033 | 2002.0 | 2007.0 |
| 0.021 | 2002.0 | 2008.0 |
| 0.029 | 2003.0 | 2004.0 |
| 0.073 | 2003.0 | 2005.0 |
| 0.027 | 2003.0 | 2006.0 |
| 0.041 | 2003.0 | 2007.0 |
| 0.007 | 2003.0 | 2008.0 |
| 0.037 | 2004.0 | 2005.0 |
| 0.008 | 2004.0 | 2006.0 |
| 0.006 | 2004.0 | 2007.0 |
| 0.012 | 2004.0 | 2008.0 |
| 0.053 | 2005.0 | 2006.0 |
| 0.044 | 2005.0 | 2007.0 |
| 0.024 | 2005.0 | 2008.0 |
| 0.005 | 2006.0 | 2007.0 |
| 0.040 | 2006.0 | 2008.0 |
| 0.038 | 2007.0 | 2008.0 |

##### 5.1.1 Degree Distributions

### 6 Comparing Louvain to Feasible Results

To assess the level of similarity of the feasible maps with the optimal regionalizations produced by the Louvain community detection algorithm, we assessed the percent match and the Normalized Mutual Information Scores (NMI) for each season using the .8 threshold, as seen in Figure S6

### 7 Comparison to HHS regions

We evaluated the epidemiological dynamics exhibited by each region of the Department of Health and Human Services regions in Figure S7. There is significant variability in the epidemic dynamics observed within each region. The peak times are often not significantly different for each season, and the HHS regions had low mutual information similarity scores to the Louvain results for each season which ranged from .07 to .24 (Table S7) suggesting that the HHS regions, and thus current disease management strategies that are based on geographic mega-regions do not capture underlying epidemiological dynamics.

### 8 Granger Casuality

To characterize the forecasting potential of epidemic dynamics of earlier regions to predict dynamics in later regions, we consider Granger causality for all pairs of regions, *i* and *j*, in which region *i* has an earlier epidemic peak than region *j*. We performed the Granger causality test on the average of all county epidemic time series for each region, and considered all weekly lags up to five weeks. We calculated p-values using the chi2 test. The directed acyclic graphs in Figure S8 summarize our results, and highlight that all pairs of ordered regions are in a Granger causal relationship. However, we note that we did not find this result for all possible lags, reducing the robustness of this result.

**Figure S6:**
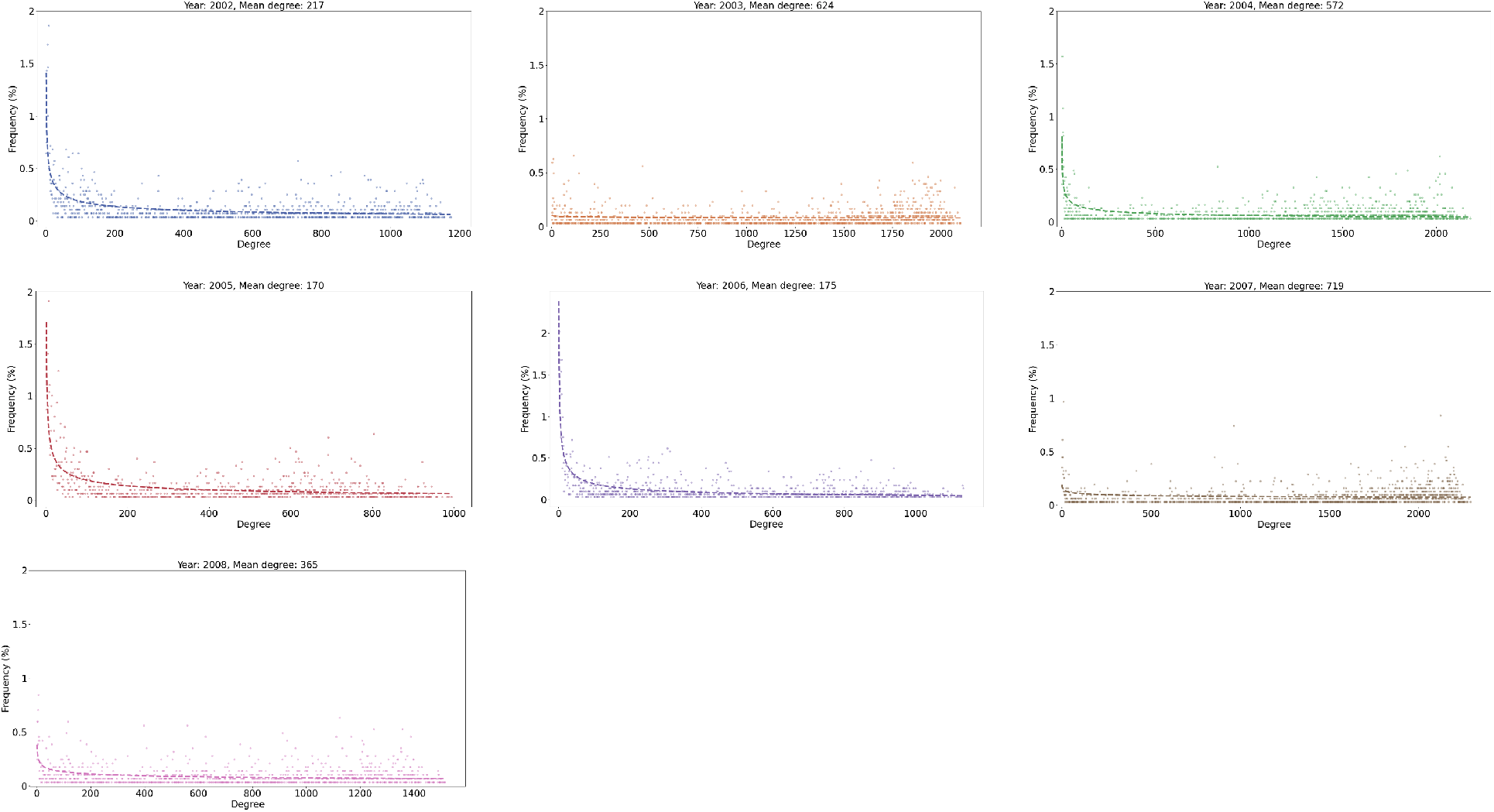
Degree Distributions Diagrams. Here we present degree distribution diagrams for each flu season using the .8 threshold. Mean degrees varied from 170 to 719 which suggests

**Table S6:**
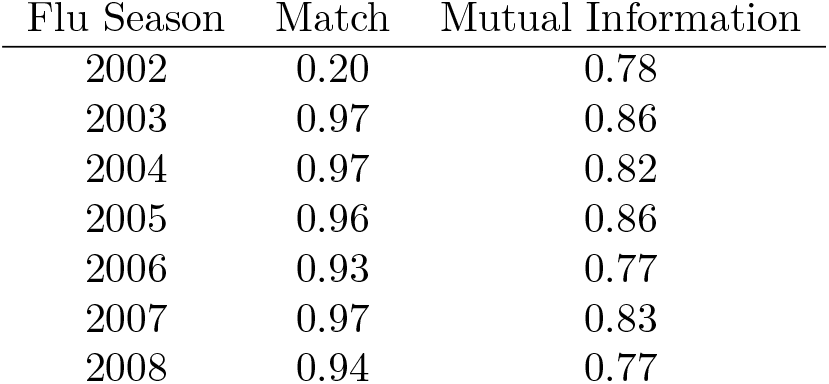
Assessing similarity of Louvain results to feasibility results. To determine the impact of increasing spatial contiguity of the regions resulting from the Louvain community detection algorithm on the complex network, we calculated the percent match and normalized mutual information score between the two for each flu season.

### 9 Random Forest Models

#### 9.1 Covariates

We considered the following continuous covariates: log population density, average household size, child and adult populations, toddler and elderly influenza vaccine coverage, population protected due to prior season exposure, percent H3 subtype among influenza type A samples, percent B type among positive influenza samples, hospitals per capita, percentage of single-person households, percentage of the population in poverty, percent of the population with health insurance, percentage of physicians reporting to the medical claims data, and all visits per capita reported to the medical claims data. Data collection and processing for these covariates has been previously described in [1]. We added three county-level covariates as potential predictors of regionalization clustering: latitude, Euclidean distance from most probable source county, and pre-epidemic anomaly in specific humidity.

**Figure S7:**
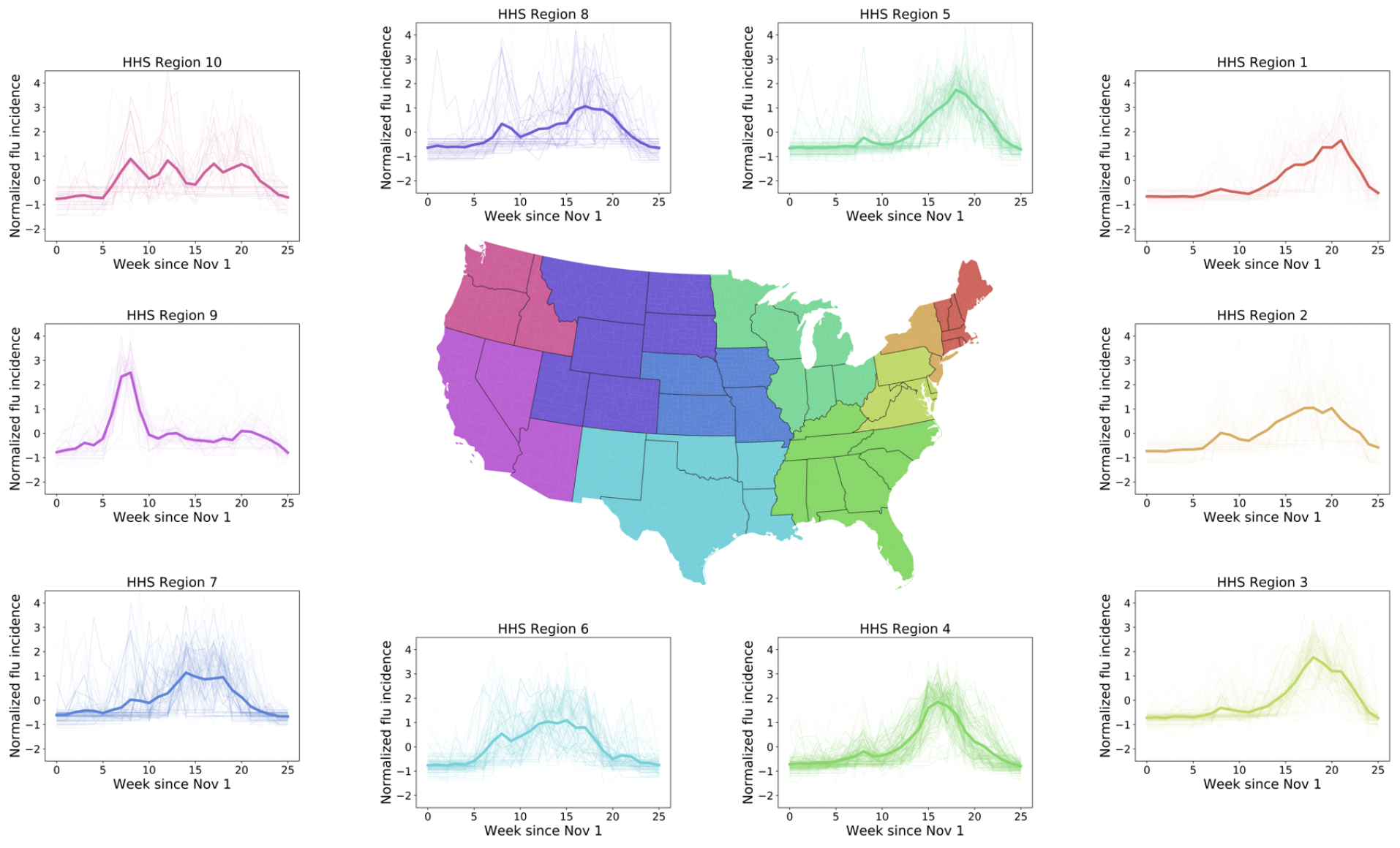
Community time series by HHS region. Here we show the epidemic time series using HHS regions as the community structure for all counties in the U.S for the 2005-2006 flu season. Most regions do not experience uniform epidemics and therefore do not experience similar influenza dynamics.

**Table S7:** Similarity between Louvain regionalization and HHS Regions. Normalized Mutual Information Scores for the regions our method identified and the corresponding HHS regions.

| Mutual Information | Flu Season |
| --- | --- |
| 0.24 | 2002.0 |
| 0.18 | 2003.0 |
| 0.07 | 2004.0 |
| 0.24 | 2005.0 |
| 0.15 | 2006.0 |
| 0.07 | 2007.0 |
| 0.11 | 2008.0 |

**Figure S8:**
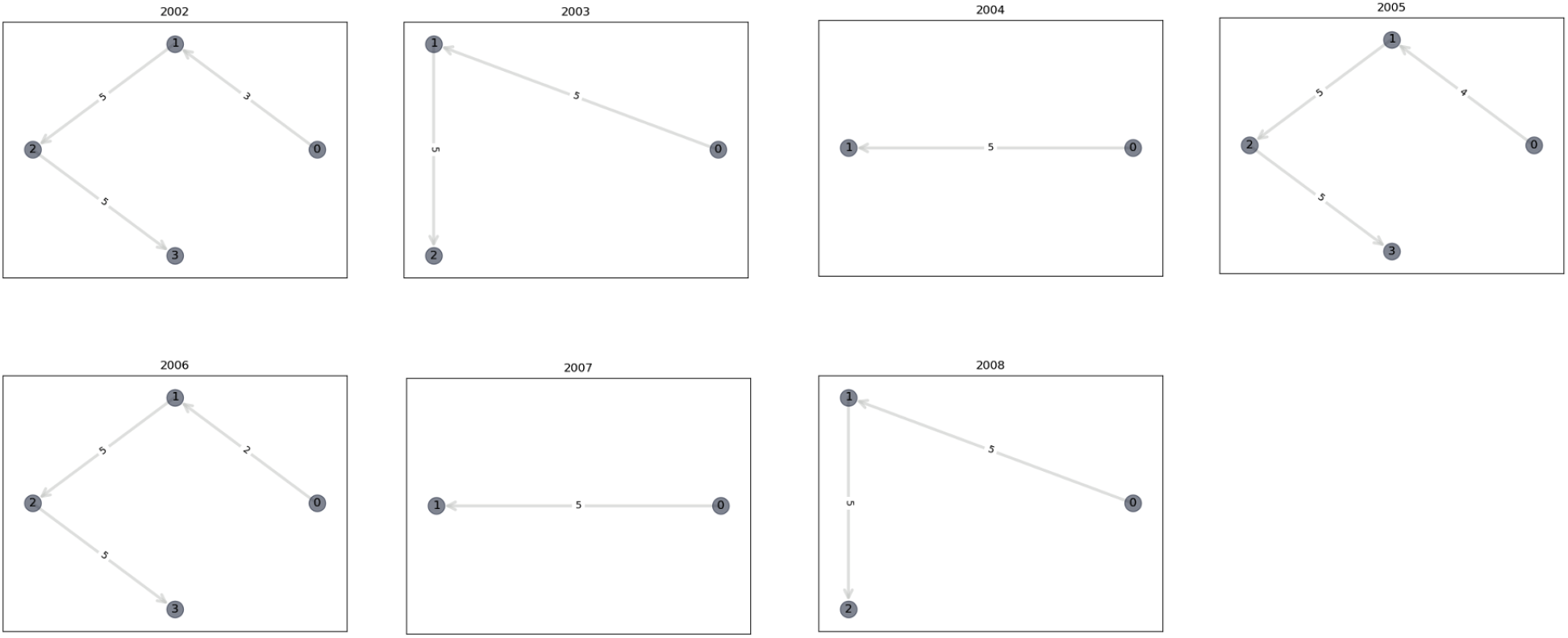
A summary of the results of the Granger causality tests. Each panel corresponds to a different influenza season, and illustrates the directed acyclic graph in which a node represents a region (with increasing region ids representing later epidemic peaks) and the directed edge from node *i* to node *j* represents that the epidemic dynamics of region *i* Granger cause the epidemic dynamics in region *j*. The edgeweight denotes the number of lags for which the Granger relationship between the nodes exists. Any edge pair not shown suggests that the test was not significant for any lags.

Latitude was added as a predictor because latitude is correlated with global influenza seasonality and peak timing and may serve as a proxy for typologies that jointly characterize influenza trends by temperature and humidity[60]; higher latitude locations tend to have annual influenza epidemics with greater peak intensity than lower latitude locations, which tend to be characterized by biannual or endemic influenza transmission.

In considering the hierarchical, wave-like spread of influenza [14], we hypothesized that regionalization clusters might be predicted by their distance from most probable source county in a given season. To identify the most probable source county, we first identified the top 10% of counties with earliest epidemic onset according to the detrended ILI ratio. Using the latitude and longitude of county centroids, we calculated the Euclidean distance between these top 10% most probable source locations and all other counties. We then used the Pearson correlation coefficient (*H*_*o*_: no difference from zero) between distance to potential source location and onset week to identify the most probable county or state source locations for a given influenza season (higher correlation coefficient means higher probability of being source location). The covariate was then presented as the Euclidean distance to the most probable source county.

Influenza season onset has previously been linked with anomalies in absolute humidity in temperate regions[61], and we hypothesized the seasonal onset might predict regionalization clusters. We calculated this county-level covariate as the mean local daily deviation in specific humidity (a measure of absolute humidity) over the four weeks prior to seasonal onset, where the deviation is the difference between daily absolute humidity and the daily average values over the study period.

#### 9.2 Results

We examined the performance of our random forest models by comparing their test and out-of-bag errors. We also compared our primary results (15% sample test set) to an analogous set with a 30% sample test set.

**Table S8:**
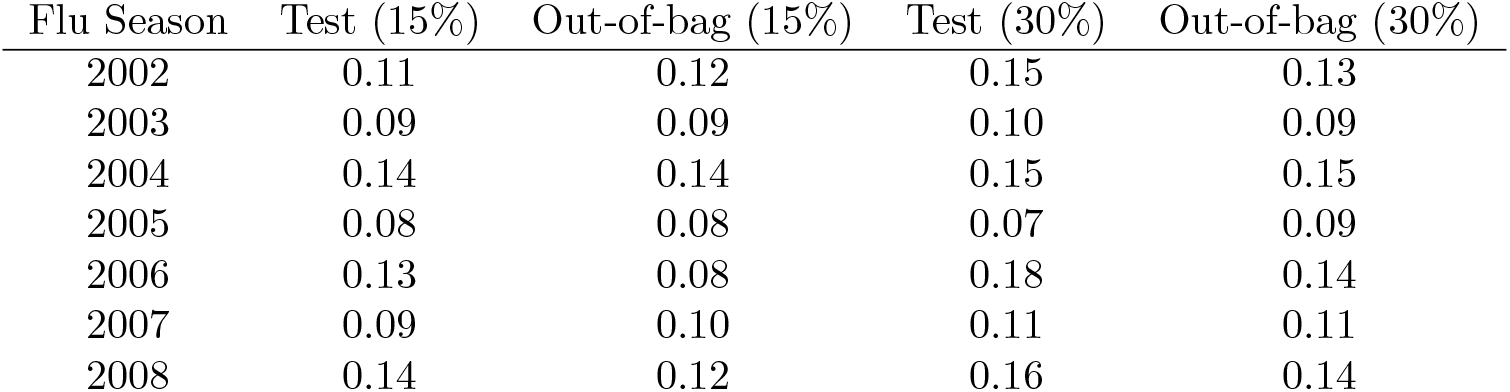
Test error and out-of-bag error for random forest models by influenza season with 15% and 30% sample test sets. These errors are reported as proportion of misclassified counties.

| Flu Season | Test (15%) | Out-of-bag (15%) | Test (30%) | Out-of-bag (30%) |
| --- | --- | --- | --- | --- |
| 2002 | 0.11 | 0.12 | 0.15 | 0.13 |
| 2003 | 0.09 | 0.09 | 0.10 | 0.09 |
| 2004 | 0.14 | 0.14 | 0.15 | 0.15 |
| 2005 | 0.08 | 0.08 | 0.07 | 0.09 |
| 2006 | 0.13 | 0.08 | 0.18 | 0.14 |
| 2007 | 0.09 | 0.10 | 0.11 | 0.11 |
| 2008 | 0.14 | 0.12 | 0.16 | 0.14 |

**Table S9:** Random forest model prediction error for one-season-ahead regionalization clusters with 15% and 30% sample test sets. These errors are reported as proportion of all county pairs that incorrectly fall in different clusters.

| Model Year | Prediction Year | Prediction Error (15%) | Prediction Error (30%) |
| --- | --- | --- | --- |
| 2002 | 2003 | 0.61 | 0.62 |
| 2003 | 2004 | 0.31 | 0.30 |
| 2004 | 2005 | 0.49 | 0.48 |
| 2005 | 2006 | 0.55 | 0.55 |
| 2006 | 2007 | 0.28 | 0.33 |
| 2007 | 2008 | 0.50 | 0.50 |

Out-of-bag errors and test errors for influenza season models ranged from 8 to 14% for the 15% sample test set results (Table S8). There were no major differences in the results between sample test set sizes, although both test errors and out-of-bag errors were slightly higher among the models with the 30% sample test set.

We used our influenza season random forest models to predict regionalization clusters for the following year’s influenza season. We then compared the similarity of the predicted clusters to the clusters that were identified by the regionalization process after the end of the real influenza season. We calculate prediction error as the proportion of counties that were incorrectly predicted to fall into different clusters (Table S9). The one-season-ahead prediction errors were large (range 28-61%) relative to the out-of-bag errors, but there was little difference in prediction performance between the 15% and 30% sample test sets.

The most important variables in the 15% sample test set were also the most important variables in the 30% sample test set, with very similar results for the mean decrease in accuracy across influenza seasons (Figure S9)

**Figure S9:**
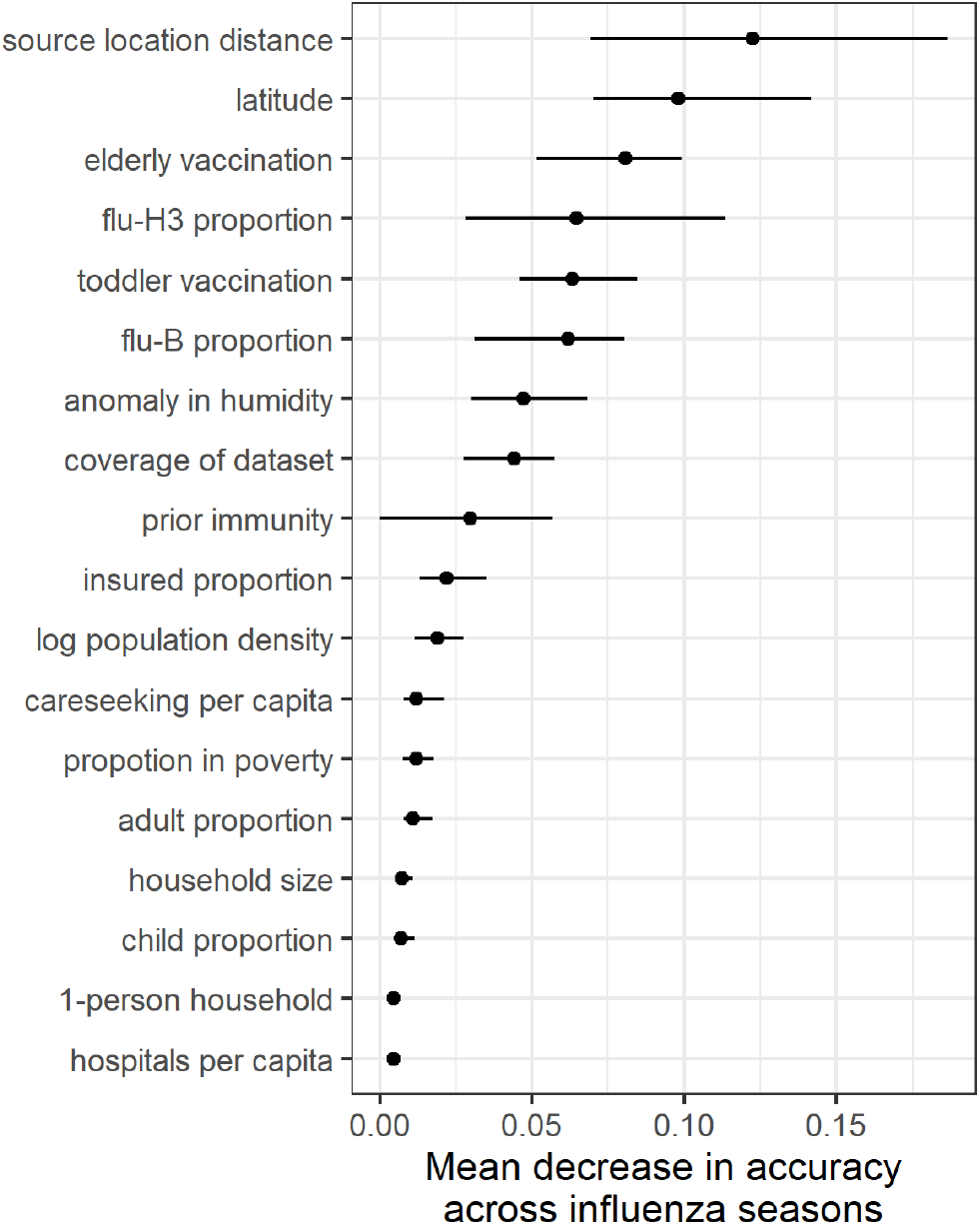
Variable importance for random forest classification of influenza season clusters using the 30% sample test set for sensitivity analysis. Variable importance is represented as the mean decrease in accuracy across influenza seasons (point is mean of the mean decrease across seasons, line range is the range of mean values across seasons).

